# High prevalence of asymptomatic and sub-patent *Plasmodium falciparum* infections in Urban Bouaké, Côte d’Ivoire, but no *hrp2* deletions: Implications for Malaria Control

**DOI:** 10.1101/2023.11.08.23298263

**Authors:** André Barembaye Sagna, Yilekal Gebre, Claudia Abigail Vera-Arias, Dipomin François Traoré, Bertin N’cho Tchekoi, Serge Brice Assi, Amnan Alphonsine Koffi, Christophe Rogier, Franck Remoue, Cristian Koepfli

## Abstract

Asymptomatic *Plasmodium falciparum* infections are common in endemic settings, yet diagnosing these infections remains challenging because they are often below the limit of detection of conventional light microscopy (LM) or rapid diagnostic tests (RDTs). Deletions of the *hrp2*/*3* gene, encoding the protein detected by the most sensitive class of RDTs, present a further threat. In this study, the prevalence of asymptomatic and sub-patent *P. falciparum* infections was characterized in the rainy season in the city of Bouaké, Central Côte d’Ivoire. A cross-sectional survey was conducted in nine neighborhoods of Bouaké, Côte d’Ivoire, in 2016. Matched LM, RDTs, and *var*ATS qPCR, were used to determine the prevalence of *P. falciparum* infections and to compare the performance of the different diagnostic methods. *P. falciparum hrp2*/*3* deletions were typed by digital PCR. Among 2313 individuals, malaria prevalence was 10.8% by microscopy, 13.0% by RDT, 35.1% by qPCR, and 37.3% (863/2313) when all three methods were combined. 96.4% (832/863) of individuals with *P. falciparum* infections did not report a recent fever. 57.1% (493/863) of infections were sub-patent. The prevalence was highly heterogeneous across the neighborhoods, ranging from 15% to 55.2%, and differed among age groups (<5 years: 17.7%, 5-14 years: 42.7%, ≥15 years: 35.9%). Compared to *var*ATS qPCR, LM and RDT had a low sensitivity of 24.3% and 34.2%, respectively, although both methods were highly specific (>96%). Sub-microscopic malaria infections were more prevalent in ≥15-year-olds (69.9%, 349/499) and in the neighborhood of Dar es Salam (75.9%, 44/58). No hrp2 deletions were observed, and two samples carried *hrp3* deletion/wild-type mixed infections. The high prevalence of asymptomatic and sub-patent infections prompts the implementation of strategies targeting these parasite reservoirs to achieve reductions in malaria burden in this high-transmission city of Côte d’Ivoire.

## Introduction

The overall number of malaria infections is undeniably much higher than the estimated 241 million clinical cases (1) in 2020, owing to the large proportion of asymptomatic infections among people living in endemic areas (2). Asymptomatic (or subclinical) malaria infection refers to malarial parasitemia of any density in the absence of fever or other acute symptoms in individuals who have not received recent antimalarial treatments (3). While some asymptomatic infections may have parasitemia levels that are detectable by light microscopy (LM) or rapid diagnostic tests (RDTs), others can only be detected by molecular methods and are termed sub-patent infections (4). Asymptomatic and sub-patent individuals remain untreated in countries where control focuses on clinical malaria cases and serve as reservoirs for onward transmission (5,6). Quantifying the extent of submicroscopic infections and the underlying true prevalence of parasites in different settings is central to understanding the dynamics of malaria transmission and adapting control measures accordingly.

The asymptomatic reservoir presents a major challenge for control, with up to 95% of transmission stemming from asymptomatic carriers (7–9). Numerous approaches are currently being trialed or implemented to shrink the asymptomatic reservoir, such as mass screening and treatment (10,11), reactive case detection (12), focal mass drug administration (13), or seasonal malaria chemoprophylaxis (14).

The success of control strategies for conducting active screening for infections relies heavily on the sensitivity of the diagnostic test used. These programs typically use RDTs, the only diagnostic method that can readily be used away from health centers. Yet, the sensitivity of RDTs is limited. Compared to highly sensitive PCR, RDTs often miss more than half of all infections (15). The most sensitive RDTs for *P. falciparum* detect the HRP2 and HRP3 proteins. Deletions of the *hrp2*/*3* genes result in false-negative RDTs, even if parasite density is high. Initially reported in Peru (16), the frequency of *hrp2*/*3* is high in the Horn of Africa (17–20), and deletions were also reported from Ghana (21,22). Surveillance of *hrp2*/*3* is needed to decide whether HRP2-based RDTs can be used for *P. falciparum* diagnosis.

The epidemiology of urban malaria in Côte d’Ivoire is not well documented, with few studies conducted in the southern (23–25) and northern parts of the country (26). These studies found a high prevalence of infection by microscopy, ranging from 12% in the north to more than 60% in the south. This difference in infection prevalence could mainly be explained by climatic factors. The climate is tropical (Sub-Sudanese) in the north with two main seasons, and equatorial in the south with two rainy seasons and two dry seasons (27). Other factors such as age groups (5-14 years), traveling to rural areas, and agriculture practices have also been found to be risk factors for malaria infections in urban areas (23–26).

Bouaké, in central Côte d’Ivoire, is the second-most populous city in the country, with more than 750,000 inhabitants in 2015. Like many cities in sub-Saharan Africa, urban agriculture is widespread within and around the city to increase food security. This agricultural activity has been shown to significantly impact malaria transmission with the creation of numerous larval habitats and vector proliferation (28–30). Bouaké is a highly endemic area with >160,000 malaria cases recorded each year (annual incidence >200 per 1000 inhabitants) and more than 100 deaths (31). Bouaké suffered from an armed conflict and a sociopolitical crisis from 2000 to 2011, leading to large population moves, environmental modifications, and interruption in the implementation and/or maintenance of malaria control strategies for the whole period (32,33).

Following the crisis, the population has reinvested in the city and resettled in formerly abandoned neighborhoods. Only a few epidemiological studies have been conducted in the city of Bouaké, and the real burden of malaria is underestimated, with no published data available on the prevalence of asymptomatic and submicroscopic malaria infections. Its characterization will help the national malaria control program (NMCP) adapt control measures for a more efficient use of limited resources.

The objectives of this study were to measure the burden of asymptomatic and sub-patent *P. falciparum* infections by LM, RDT, and qPCR in the highly endemic city of Bouaké and to investigate potential *P. falciparum hrp2* and/or *hrp3* deletion.

## Materials and Methods

### Ethical approval and source of funding

Funding was provided by the French Initiative 5% and Expertise France, Grant N°12INI210. This study followed the ethical principles recommended by the Edinburgh revision of the Declaration of Helsinki and was approved by the Ethics Committee of the Ministry of Health of Côte d’Ivoire (June 2015; No. 029/MSLS/CNER-dkn) and the University of Notre Dame Institutional Review Board (18-08-4803). Before collecting samples, all participants over the age of 18 provided written informed consent, as did the parent/guardian of each participant under the age of 18.

### Study area and demographic data collection

In Côte d’Ivoire, malaria transmission occurs all year round, with a peak during the rainy season. The annual incidence rates vary between 100 and >500 cases per 1000 inhabitants, with more than 80% of the whole Ivoirian population living in areas with an annual incidence rate ≥300 per 1000 inhabitants (31). Despite a national coverage of long-lasting insecticidal nets (LLINs) estimated at 95% since 2015, the number of malaria-related cases has been increasing since 2016 from 3,754,504 to 7,295,068 cases in 2021 (34). The number of malaria-related deaths has decreased during the same period, from 3,340 to 1,276 deaths.

The survey was conducted in three health districts of the city of Bouaké: Northwestern-Bouaké, Northeastern-Bouaké, and Southern-Bouaké. All health districts had a high coverage rate (>80%) of standard pyrethroid-based LLINs. This area has year-round malaria transmission, with a peak during the wet season (April–October). *P. falciparum* is the dominant species identified in more than 95% of malaria cases (35). The *Anopheles gambiae* species complex is the dominant vector species, with *An. funestus* and *An. nili* also present. The local vector populations are highly resistant to almost all classes of insecticides used for vector control (36,37).

A cross-sectional study was performed from August 1^st^ to August 27^th^, 2016, in nine neighborhoods of the city (three neighborhoods per health district): Dar-es-Salam, Djezoukouamekro, and N’gattakro in Northwestern-Bouaké; Sokoura, Belle-ville, and Attienkro in Northeastern-Bouaké; Kennedy, Air-France, and N’gouattanoukro in Southern-Bouaké. This study was part of a large operational research project named PALEVALUT (http://www.mesamalaria.org/mesa-track/palevalut), which aimed to evaluate the effectiveness of malaria control resources currently deployed and identify the barriers to their effectiveness in five different sub-Saharan countries (Benin, Cameroon, Côte d’Ivoire, Niger, and Madagascar). In each selected neighborhood, 50 households were randomly selected, and all members who were at home were invited to participate in the study. After enrollment, all households were geo-located using GPS (Garmin, Switzerland) instruments with up to 5 m of accuracy. A socio-demographic questionnaire was completed with the head of the family to collect information about the sanitary conditions of the household and household characteristics (house type and construction materials, bathroom type, access to public services, etc.). Information on age, sex, history of fever, LLIN use, and knowledge of malaria was recorded for each participant.

### Case definitions

An asymptomatic malaria infection was defined as a *P. falciparum* positive test result in an afebrile individual (temperature ≤37.5°C) at the time of sampling with no history of fever in the previous 48 hours. A clinical malaria case was defined as a positive test result with a history of fever or a temperature >37.5 °C. Sub-patent infections were defined as infections that were only positive by *var*ATS qPCR.

### Blood sample collection

During the house visit, blood samples were collected from each study participant using a 1 mL insulin syringe (Luer-Lok™, BD, Franklin Lakes, NJ, USA) to prepare thick and thin blood smears (both thick and thin smears were collected on the same slide), one RDT, and two spots on Whatman qualitative filter paper N°3. The remaining blood was poured into a 1.5-mL microcentrifuge tube. At the laboratory, microcentrifuge tubes were centrifuged, and plasma was removed and stored at -20°C. The red cell pellets were also stored at -20°C until DNA extraction. All samples of each study participant were labeled with a unique code and stored accordingly until use.

### RDT-based diagnosis and treatment

The Pf/HRP2-based RDT (Standard Diagnostics Inc.; Yongin, Republic of Korea) recommended by the NMCP of Côte d’Ivoire was used in this study. This RDT is based on histidine-rich protein 2 (HRP2) detection and is specific to *P. falciparum* (Pf). All febrile individuals that tested positive for RDT were treated with anti-malarials according to the national drug policy. Those found positive by microscopy or qPCR were not treated because microscopy examination and qPCR assays were not performed at the point of collection.

### Microscopic examination

Giemsa-stained peripheral blood smears were examined under a compound microscope (Olympus CX21 LED) at 1000× magnification to detect malaria parasites by trained technicians. Ring, trophozoite, or gametocyte stages were identified and assessed for parasite density (parasites/µl blood) against 200 white blood cells (WBCs), and densities were calculated considering an average of 8000 WBC/µl.

All positive slides and 10% of randomly chosen negative blood slides were cross-examined by independent laboratory personnel as part of a standard protocol to monitor examination quality. Microscopists who examined the participants’ blood slides were not aware of the RDT results.

### DNA extraction and qPCR detection of P. falciparum infections

DNA was extracted from 100 μL of blood using the Genomic DNA Extraction Kit (Macherey-Nagel, Düren, Germany) and eluted in 100 µL of elution buffer. 4 µL of extracted DNA was screened for *P. falciparum* using ultrasensitive qPCR that amplifies a conserved region of the var gene acidic terminal sequence (*var*ATS) according to a previously published protocol (38). The qPCR results were converted to parasites/μL using an external standard curve of ten-fold serial dilutions (5-steps) of 3D7 *P. falciparum* parasites quantified by droplet digital PCR (ddPCR) (39,40).

### P. falciparum hrp2/3 typing by digital PCR

Samples positive for *P. falciparum* at a density of approximately >5 parasites/μL by *var*ATS qPCR were typed for *hrp2* and *hrp3* deletion by dPCR (41). To this end, the assay was successfully transferred from the BioRad QX 200 droplet digital PCR system to the Qiagen QIAcuity dPCR system. Assay conditions were maintained as previously published (41). In this assay, *hrp2* or *hrp3* and a control gene (*serine-tRNA ligase*) were quantified in a single tube with very high specificity, thus providing highly accurate data on deletion status. The assay amplified parts of *hrp2* exon 2 that encode for the antigen detected by the RDT.

### Data analysis

Field data were collected and stored using the Open Data Kit toolkit. Electronic data was exported to MS Excel for analysis. A data summary was done using descriptive statistics. Malaria prevalence was calculated for each diagnostic test (light microscopy and RDT) and compared to the gold standard of *var*ATS qPCR using McNemar’s or Cochran’s test. The comparison of malaria prevalence by age group, gender, neighborhood, symptoms, and LLIN use was done using the Pearson χ2 test or Fisher’s exact test, as appropriate. Differences in parasite densities between age groups were determined by ANOVA’s Dunn’s multiple comparisons test. The limits of detection (LoD) of light microscopy (LM) and RDT, defined as the lowest parasite density that would be detected with 95% probability, were determined by logistic regression based on *var*ATS qPCR parasite density. Univariate and multivariate logistic regression models were used to investigate the association between sub-patent malaria infections and risk factors.

Variables significant at a p-value of 0.30 in the univariate logistic regression were selected for the multivariate logistic regression analysis model. For all statistical tests, a 5% level of significance was used. Odds ratios with 95% confidence intervals (CI) were reported. All statistical analyses were performed using Graph Pad Prism 9.5 (Graph Pad Software, San Diego, CA, USA) and MedCalc 20.218 (MedCalc Software Ltd., Ostend, Belgium).

## Results

### Study population demography

Matched LM, RDT, and varATS qPCR data were available for 2313 individuals (Figure 1). The median age was 18 years (interquartile range: 8-33). Females represented 57.2% of the study population. The majority of individuals (60.4%) reported using an LLIN within the last 15 days, which varied between neighborhoods (Table 1).

**Table 1:**
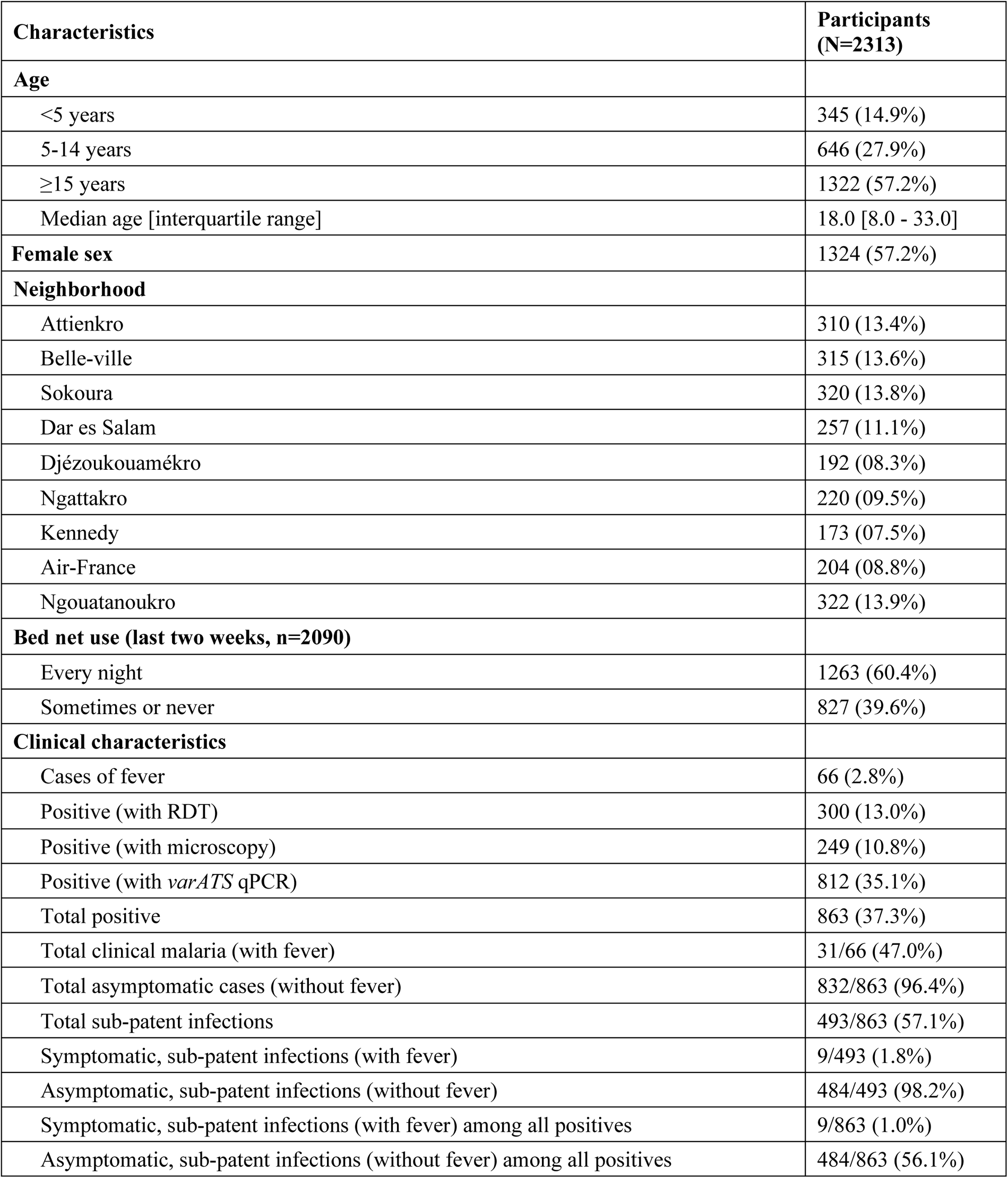
Social demographic and clinical characteristics of the study population: Bouaké, Côte d’Ivoire, 2016.

**Figure 1:**
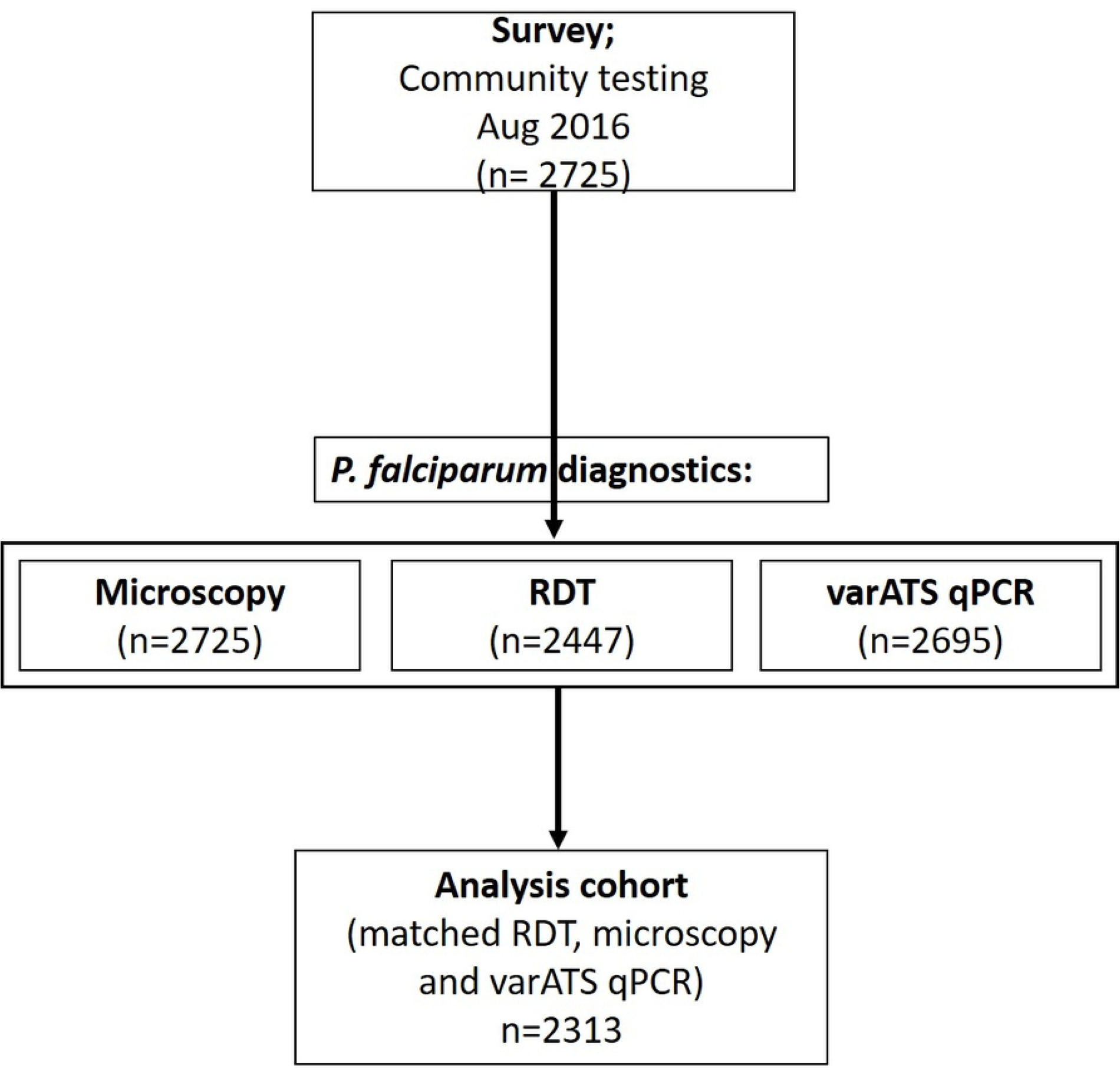
Study workflow diagram. Data for analysis consists of participants for whom results from all three tests were available.

### Symptomatic, asymptomatic and sub-patent P. falciparum infections

Among the full study population, 37.3% (863/2313) of participants were diagnosed positive by LM, RDT, and qPCR combined (**Table 1**). **Figure 2** shows the distribution of *P. falciparum* infections according to the three diagnostic methods used. The prevalence of *P. falciparum* infections varied significantly according to the diagnostic method and was 10.8% (249/2313) by LM, 13% (300/2313) by RDT, and 35.1% (812/2313) by qPCR (Cochran’s Q= 823.9, DF= 2, p<0.001). Of all *P. falciparum* infections, 96.4% (832/863) were classified as asymptomatic and 57.1% (493/863) as sub-patent (**Table 1**, **Figure 2**). Sixty-six (66) persons reported fever or a history of fever within 48 hours, of which 31 (47%) were positive by LM, RDT, and qPCR combined. The number of clinical malaria cases was variable depending on the diagnostic method and was 17 by LM, 20 by RDT, and 27 by qPCR. Nine clinical malaria cases were missed by both LM and RDT (**Figure 2**).

**Figure 2:**
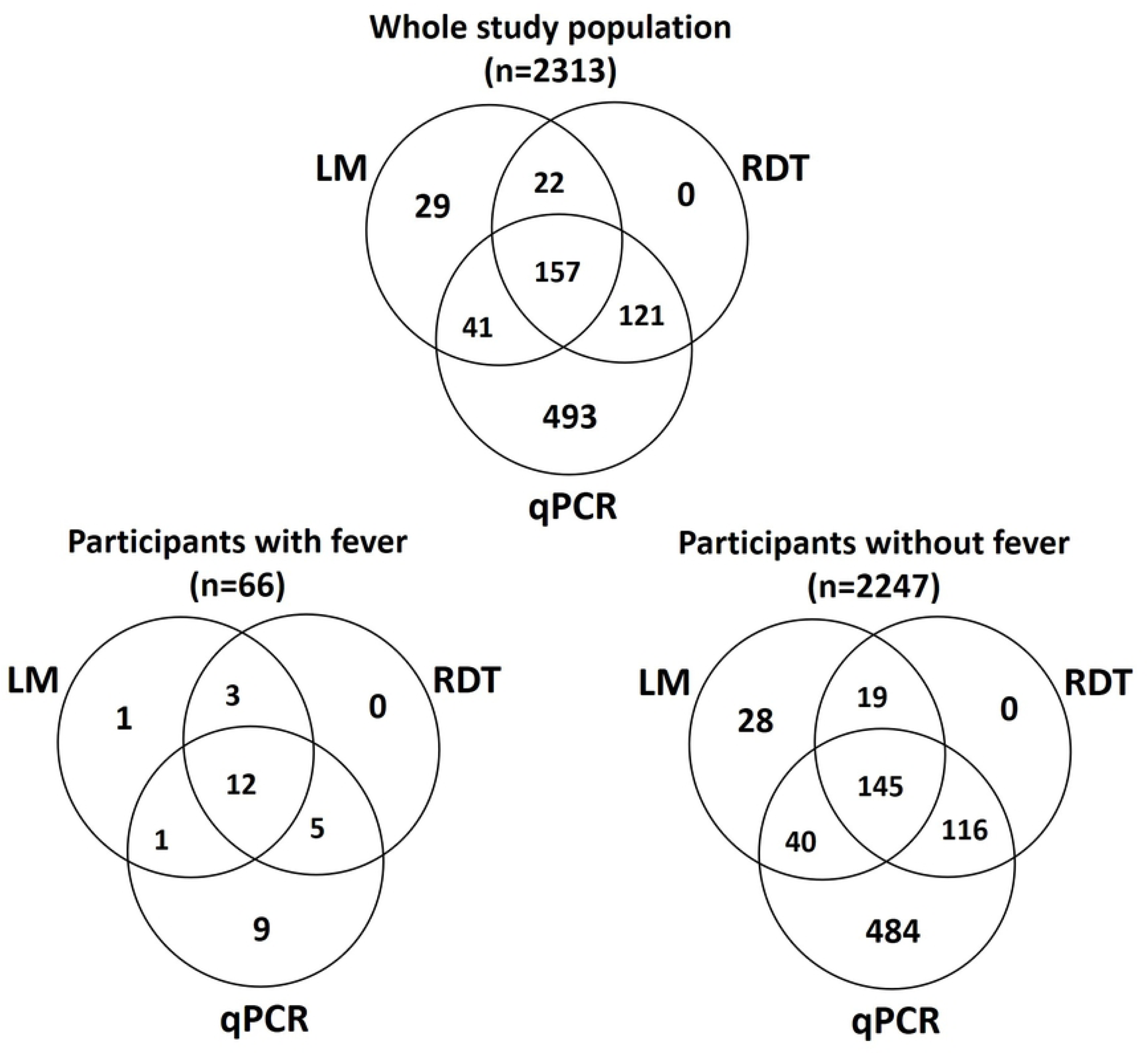
Distribution of *P. falciparum* infections by the three diagnostic methods.

*P. falciparum* infection prevalence differed significantly among age groups, sex, neighborhoods, and symptoms (**Table 2**). The prevalence of infections peaked in 5–14-year-olds (irrespective of diagnostic method, all p< 0.0001). Although malaria prevalence tended to be lower in individuals ≥15 years compared to <5 years by LM and RDT, it was 2-fold higher in individuals ≥15 years compared to <5 years by qPCR. Prevalence was higher in males compared to females by all three diagnostic methods (all p< 0.05; **Table 2**). The prevalence was highest in Attienkro, health district of Northwestern Bouaké (28.7% by LM, 19.0% by RDT, and 55.2% by qPCR). The lowest prevalence by LM (1.6%) and qPCR (15.0%) was recorded in Sokoura, health district of Northwestern Bouaké, and by RDT (5.1%) in Dar es Salam, health district of Northeastern Bouaké (**Table 2**). Aside from the neighborhood of Attienkro, health district of Northeastern Bouaké, where prevalence was higher for LM (28.7%) compared to RDT (19.0%, p< 0.001), all neighborhoods displayed an equal or higher prevalence by RDT as compared to LM. qPCR detected higher numbers of infections than LM or RDT in all neighborhoods (all p< 0.001).

The prevalence of *P. falciparum* infections was significantly higher in individuals who reported fever or a history of fever within 48 hours compared to afebrile individuals by LM and RDT (all p< 0.0001), but not by qPCR (p= 0.316).

The overall parasite density expressed as geometric mean parasite density by qPCR was 40 parasites/µL (95% CI: 33.7-47.5, **Figure 3A**). Individuals who were positive by all three tests had a significantly higher parasite density (856.8 parasites/µL; 95% CI: 707.3–1,038) than those who were positive by *var*ATS qPCR only (13.7 parasites/µL; 95% CI: 11.4–16.3) (p< 0.0001) (**Figure 3A**). Parasite density varied significantly by age group and was lower in ≥15-year-olds (27.2 parasites/µL; 95% CI: 22.2-33.5) compared to <5-year-olds (73.5 parasites/µL; 95% CI: 36.9-146.2; p=0.004) and 5-14-year-olds (76.5 parasites/µL; 95% CI: 55.9-104.7; p< 0.001, **Figure 3B**).

**Table 2:** Prevalence of *P. falciparum* infections by microscopy, RDT and *var*ATS qPCR (N=2313).

| Characteristics |  | N | Tested positive by LM | *p*-value | Tested positive by RDT | *p*-value | Tested positive by qPCR | *p*-value |
| --- | --- | --- | --- | --- | --- | --- | --- | --- |
| Age group | <5 | 345 | 30 (8.7%) | **<0.0001** | 42 (12.2%) | **<0.0001** | 61 (17.7%) | **<0.0001** |
|  | 5-14 | 646 | 118 (18.3%) |  | 153 (23.7%) |  | 276 (42.7%) |  |
|  | ≥15 | 1322 | 101 (7.6%) |  | 105 (7.9%) |  | 475 (35.9%) |  |
| Gender | Male | 989 | 123 (12.4%) | **0.026** | 148 (15.0%) | **0.014** | 376 (38.0%) | **0.012** |
|  | Female | 1324 | 126 (9.5%) |  | 152 (11.5%) |  | 436 (32.9%) |  |
| Neighborhood | Attienkro | 310 | 89 (28.7%) | **<0.0001** | 59 (19.0%) | **<0.0001** | 171 (55.2%) | **<0.0001** |
|  | Belle-ville | 315 | 29 (9.2%) |  | 39 (12.4%) |  | 103 (32.7%) |  |
|  | Sokoura | 320 | 5 (1.6%) |  | 16 (8.4%) |  | 48 (15.0%) |  |
|  | Dar es Salam | 257 | 9 (3.5%) |  | 13 (5.1%) |  | 58 (22.6%) |  |
|  | Djézoukouamékro | 192 | 27 (14.1%) |  | 36 (18.8%) |  | 94 (49.0%) |  |
|  | Ngattakro | 220 | 16 (7.3%) |  | 17 (7.7%) |  | 65 (29.5%) |  |
|  | Kennedy | 173 | 13 (7.5%) |  | 26 (15.0%) |  | 69 (39.9%) |  |
|  | Air-France | 204 | 22 (10.8%) |  | 36 (17.6%) |  | 81 (39.7%) |  |
|  | Ngouatanoukro | 322 | 39 (12.1%) |  | 58 (18.0%) |  | 123 (38.2%) |  |
| Symptoms | Febrile | 66 | 17 (25.8%) | **<0.0001** | 20 (30.3%) | **<0.0001** | 27 (40.9%) | 0.316 |
|  | Afebrile | 2247 | 232 (10.3%) |  | 280 (12.5%) |  | 785 (34.9%) |  |
| Bed net use (last two weeks) | Every night | 1263 | 147 (11.6%) | 0.378 | 166 (13.1%) | 0.854 | 460 (36.4%) | 0.727 |
|  | Sometimes/never | 827 | 86 (10.4%) |  | 111 (13.4%) |  | 295 (35.7%) |  |

**Figure 3:**
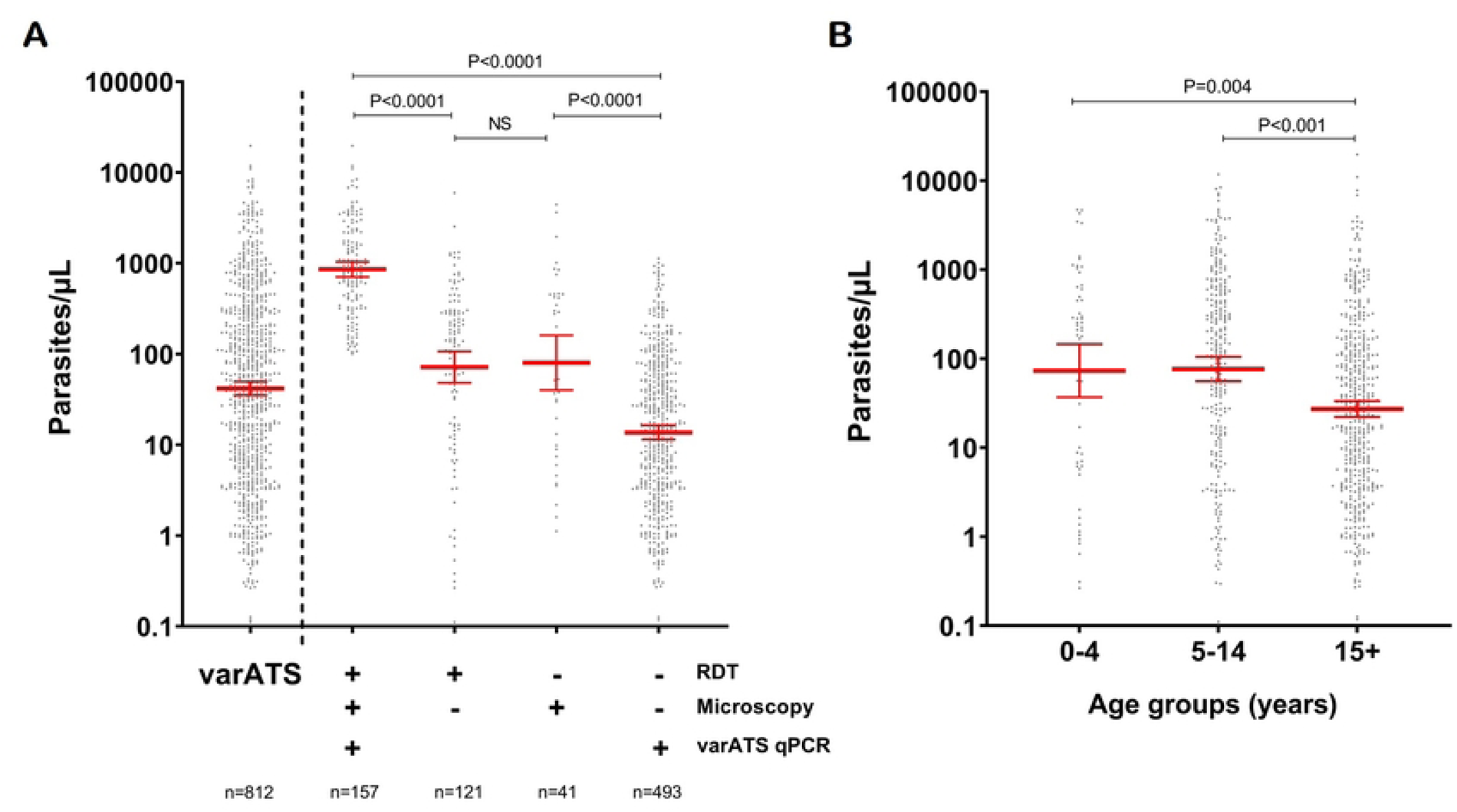
*P. falciparum* density as determined by *var*ATS qPCR. **Panel A** shows *P. falciparum* density in the whole study population and in samples tested positive (+) or negative (−) by microscopy and/or RDT. **Panel B** shows *P. falciparum* density by age group. Dots represent the parasite density of each participant. Red bars indicate geometric means with 95% confidence intervals. P-values from Dunn’s test with Bonferroni adjustment for multiple pairwise comparisons are given.

### Specificity and sensitivity of microscopy and RDT

Compared to the gold standard qPCR, the sensitivity of RDT and LM was low at 34.24% (95% CI: 30.97–37.61) and 24.34% (95% CI: 21.47–27.49), respectively (**Table 3**). However, both methods were highly specific: 98.53% (95% CI: 97.79–99.08) for RDT, and 96.60% (95% CI: 95.56–97.46) for LM. The measurement of agreement (Kappa) was high between RDT and microscopy (60.6%, p< 0.001).

**Table 3:**
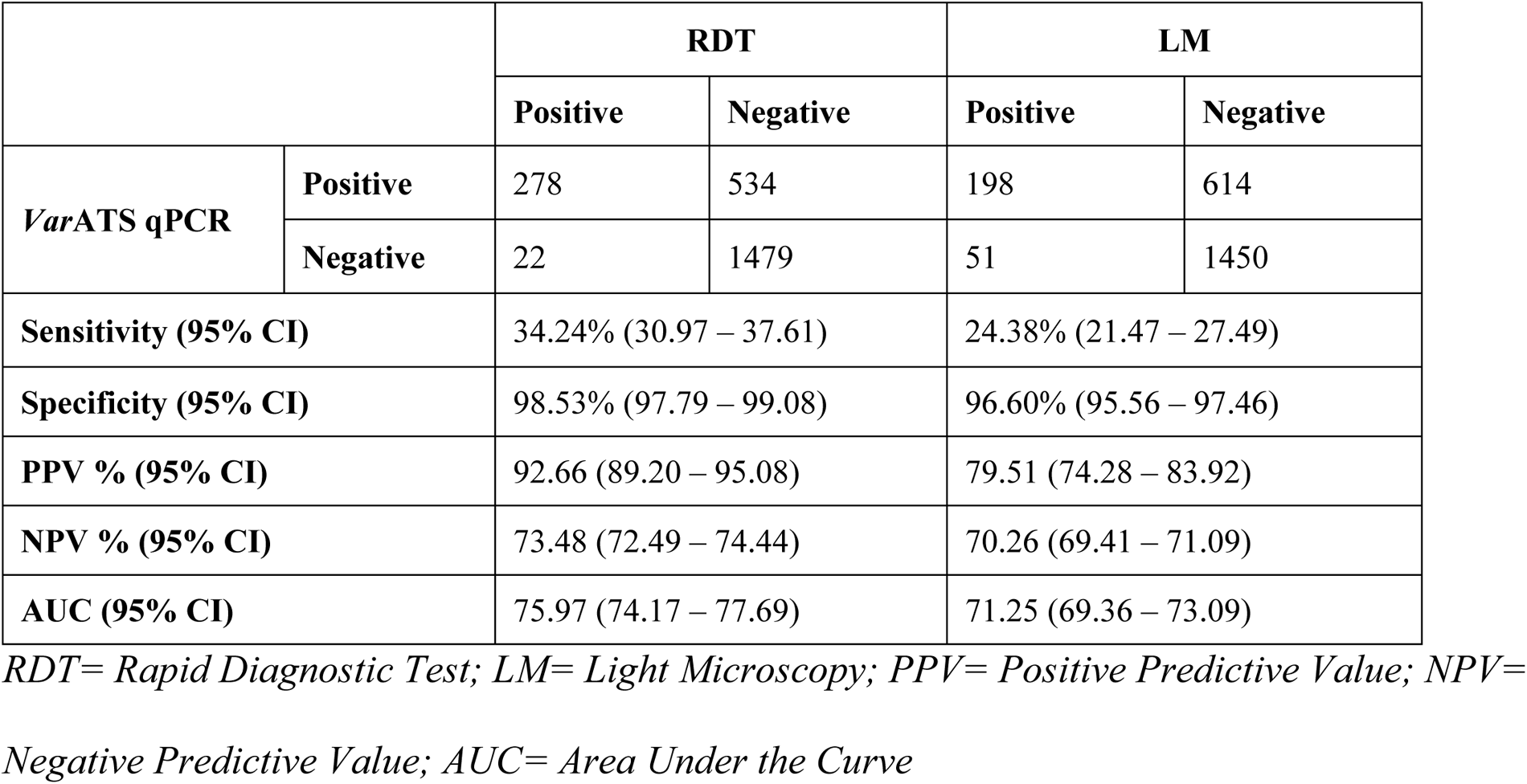
Test performance of the different diagnostic assays used for evaluating *P. falciparum* infection.

|  |  | RDT |  | LM |  |
| --- | --- | --- | --- | --- | --- |
|  |  | Positive | Negative | Positive | Negative |
| *Var*ATS qPCR | Positive | 278 | 534 | 198 | 614 |
|  | Negative | 22 | 1479 | 51 | 1450 |
| Sensitivity (95% CI) |  | 34.24% (30.97 – 37.61) |  | 24.38% (21.47 – 27.49) |  |
| Specificity (95% CI) |  | 98.53% (97.79 – 99.08) |  | 96.60% (95.56 – 97.46) |  |
| PPV % (95% CI) |  | 92.66 (89.20 – 95.08) |  | 79.51 (74.28 – 83.92) |  |
| NPV % (95% CI) |  | 73.48 (72.49 – 74.44) |  | 70.26 (69.41 – 71.09) |  |
| AUC (95% CI) |  | 75.97 (74.17 – 77.69) |  | 71.25 (69.36 – 73.09) |  |

*RDT= Rapid Diagnostic Test; LM= Light Microscopy; PPV= Positive Predictive Value; NPV= Negative Predictive Value; AUC= Area Under the Curve*

The limit of detection (LoD), which is defined here as the lowest parasite density that could be detected with 95% probability, was 1,117 (95% CI: 968–1331) parasites/µL for LM and 972 (95% CI: 834–1173) parasites/µL for RDT. The LoD of the *var*ATS qPCR method was previously determined at 0.29 (95% CI: 0.14–2.30) parasites/µL (42).

### Risk factors for sub-patent infections

Age group, the neighborhood of residency, and absence of fever within 48 hours and at the time of sampling were associated with an increased risk of sub-patent malaria infection by univariate analysis (**Table 4**). All risk factors were tested in the univariate analysis, and only those with p≤0.30, such as age group, gender, neighborhood of residency, and symptoms, were selected and included in the multivariate analysis. Only age ≥15 years (aOR: 1.09; 95% CI: 1.06 – 1.13; p< 0.0001), the neighborhood of residency, specifically Dar es Salam (aOR: 3.97; 95% CI: 1.91 – 8.27; p< 0.001), and absence of symptoms (aOR: 2.83; 95% CI: 1.22 – 6.51; p= 0.015), were significantly associated with the prevalence of sub-patent infections in multivariable logistic regression analysis. The gender and bed net use did not correlate with sub-patent malaria infection prevalence (all p> 0.05) (**Table 4**).

**Table 4:** Prevalence and risk factors for sub-patent malaria infections in malaria infected individuals (N=863)

| Risk factor | Category | Malaria positive (n=863) | Sub-patent malaria (%) | Univariate cOR [95% CI] | *p-value* | Multivariate aOR [95% CI] | *p-value* |
| --- | --- | --- | --- | --- | --- | --- | --- |
| Age group | <5 | 75 | 27 (36) | 1 |  | 1 |  |
|  | 5-14 | 289 | 117 (40.5) | 1.21 [0.71-2.05] | 0.479 | 1.0 [0.90-1.12] | 0.922 |
|  | ≥15 | 499 | 349 (69.9) | 4.13 [2.49-6.88] | **<0.0001** | 1.09 [1.06-1.13] | **<0.0001** |
| Gender | Male | 402 | 218 (54.2) | 1 | 0.108 | 1 | 0.294 |
|  | Female | 461 | 275 (59.7) | 1.25 [0.95-1.64] |  | 1.17 [0.87-1.55] |  |
| Neighborhood | Attienkro | 202 | 95 (47.0) | 1 |  | 1 |  |
|  | Belle-ville | 105 | 63 (60.0) | 1.69 [1.05-2.73] | **0.032** | 1.19 [1.06-1.33] | **0.002** |
|  | Sokoura | 50 | 31 (62.0) | 1.84 [0.97-3.47] | 0.060 | 1.27 [1.06-1.52] | **0.007** |
|  | Dar es Salam | 58 | 44 (75.9) | 3.54 [1.83-6.86] | **<0.001** | 3.97 [1.91-8.27] | **<0.001** |
|  | Djézoukouamékro | 96 | 56 (58.3) | 1.58 [0.97-2.58] | **0.069** | 1.28 [0.98-1.68] | **0.074** |
|  | Ngattakro | 65 | 44 (67.7) | 2.36 [1.31-4.25] | **0.004** | 1.41 [1.13-1.76] | **0.002** |
|  | Kennedy | 72 | 46 (63.9) | 2.00 [1.14-3.47] | **0.015** | 1.15 [1.05-1.26] | **0.004** |
|  | Air-France | 86 | 47 (54.7) | 1.36 [0.82-2.25] | 0.237 | 1.10 [1.02-1.19] | **0.012** |
|  | Ngouatanoukro | 129 | 67 (51.9) | 1.22 [0.78-1.89] | 0.384 | 1.06 [1.00-1.12] | **0.037** |
| Symptoms | Febrile | 31 | 9 (29.0) | 1 | **0.003** | 1 | **0.015** |
|  | Afebrile | 832 | 484 (58.2) | 3.34 [1.5-7.3] |  | 2.83 [1.22-6.51] |  |
| Bed net use (last two weeks) | Every night | 489 | 277 (56.6) | 1 | 0.870 | 1 | 0.430 |
|  | Sometimes/never | 311 | 178 (57.2) | 1.02 [0.77-1.37] |  | 1.13 [0.84-1.53] |  |

cOR: Crude odds ratio; aOR: Adjusted odds ratio; CI: Confidence interval;

### *P. falciparum* hrp2/3 deletion typing

A total of 344 *P. falciparum*-positive samples were successfully typed for *hrp2* and *hrp3* deletion. No *hrp2* deletions were observed. Two samples carried *hrp3* deletion/wild-type mixed infections. The first was from an 8-year-old asymptomatic male living in the neighborhood of Air France (health district of Southern Bouaké) who was negative by RDT but positive by LM and qPCR, with a parasite density of 149 parasites/µL. The second was from a 9-year-old asymptomatic female from the neighborhood of Ngattakro (health district of Northeastern Bouaké) and was also positive by LM and qPCR only, with a parasite density of 190 parasites/µL.

## Discussion

In 2016, in Bouaké, approximately a third of apparently healthy city dwellers had an asymptomatic malaria infection during the rainy (high transmission) season. More than half of malaria infections were detectable only by highly sensitive PCR (sub-patent infections).

Microscopy and rapid diagnostic tests detected fewer than 40% of qPCR-positive infections. While prevalence was high, it was lower than in 2014, when a study conducted in three neighborhoods reported a malaria prevalence of around 80% by microscopy (35). This sharp fall in malaria prevalence between 2014 and 2016 could be attributed to intensive malaria control efforts by the NMCP after the end of the crisis that occurred from 2000 to 2011. Two rounds of large-scale distribution of LLINs were done in 2011 and 2014, leading to a coverage and usage rate of 95% and 68%, respectively (43). In addition, the wide availability of RDTs and artemisinin-combination therapy (ACTs) for testing and treating symptomatic individuals helped decrease the burden of malaria in Bouaké.

The prevalence of malaria infection was highly heterogeneous across the neighborhoods, ranging from 15% to more than 55%. Likewise, the prevalence of sub-patent infections also varied across neighborhoods. The ecological environment and *Anopheles* density variability between neighborhoods could be considerable factors that explain this variation. Bouaké is characterized by the presence of lowlands used for agricultural activities, depending on the neighborhood (44). This agricultural activity has been shown to have a significant impact on vector diversity and density (30) and consequently on malaria transmission (28,29), which is highly heterogeneous across neighborhoods.

The prevalence of malaria infection was higher in school-age children (5-14 years) compared to younger ones, in line with previous studies (26,45). The high number of malaria infections detected in school-aged children raises concern because they serve as a source of onward parasite transmission (46,47), as this age group has been shown to be the least likely to benefit from universal malaria interventions such as bed nets and access to prompt diagnosis and treatment (48,49). Implementation of strategies that specifically target this age group could help further decrease malaria transmission in this highly endemic area and improve the health of schoolchildren (50–52). The proportion of sub-patent infections was higher in individuals above 15 years of age, probably because of greater immunity in older children and adults induced by cumulative exposure to the parasite (53).

The sensitivity of LM and RDT is low in areas where a substantial number of asymptomatic and low-density infections are present (54). In this study, the sensitivity of LM and RDTs was 24% and 34%, respectively, as compared to the *var*ATS qPCR. Highly sensitive RDTs (55) or loop-mediated isothermal amplification (LAMP) (56) could be promising options to improve the diagnosis of asymptomatic and sub-patent infections in areas where they are common. In Northern Côte d’Ivoire, for example, LAMP has been shown to be a relevant field diagnostic alternative to standard RDTs for identifying asymptomatic infections with a sensibility and specificity of 93.3% (95% CI 85.7–100) and 95.4% (95% CI 92.2–100), respectively (56).

For the first time in Côte d’Ivoire, parasites were typed for *hrp2*/*3* deletions. No *hrp2* deletions were detected in the study population, so the use of Pf/*hrp2*-based RDTs for the diagnosis of *P. falciparum* is still recommended. Two samples carried an *hrp3*/wild-type mixed infection. *Hrp3* deletion is not a threat for HRP2-based RDTs as long as *hrp2* is present (57). Future monitoring of *hrp2*/*hrp3* deletion is warranted in samples that are positive by microscopy or LDH-based RDT, but negative by HRP2-based RDT.

## Conclusion

The prevalence of *P. falciparum* infection was underestimated when microscopy and RDT were used for diagnosis because a substantial proportion of asymptomatic and sub-patent malaria carriers were present in the population living in the city of Bouaké and could only be diagnosed using the ultra-sensitive PCR diagnostic method (*var*ATS qPCR). The prevalence of malaria infection was highly heterogeneous across the city, and older children and adults constitute the reservoirs of the parasite. These hidden parasite reservoirs are a major challenge for the malaria control program in the city. Various strategies, such as intermittent preventive treatment, periodic mass parasite screening and treatment, or mass drug administration, could be suggested to further reduce the burden of malaria in this city. But before that, more follow-up cohort studies will be needed to identify the role of asymptomatic cases in the dynamics of malaria transmission, its seasonal variability, and its contribution to the incidence of malaria in the region, and to provide support in planning an effective and appropriate malaria elimination strategy to implement.

## Data Availability

The data can be obtained from the corresponding authors.

## Acknowledgements

We gratefully acknowledge the populations of 9 neighborhoods (DAR, DJZ, NGA, SOK, BLV, ATK, KEN, AIF, and NGO) of Bouaké, especially householders, housewives, guardians of children, and all the technical staff involved in blood sample collection in the field, for their kind support and collaboration. We would also like to thank Mahamadou Barro, PhD student in mathematics, for his invaluable help in analyzing the data.

## Authors’ contributions

ABS, CK and FR conceived the study, ABS, DFT, SBA, TBN conducted field work and recorded demographic data; YG and CAVA conducted the lab work, ABS analyzed the data and wrote the manuscript. AAK, CR and CK revised the manuscript. All authors read and approved the final manuscript.

